# Preliminary evaluation of COVID-19 disease outcomes, test capacities and management approaches among African countries

**DOI:** 10.1101/2020.05.16.20103838

**Authors:** Adebayo A. Otitoloju, Esther O. Oluwole, Kafilat A. Bawa-Allah, Mayowa J. Fasona, Ifeoma P. Okafor, Chukwuemeka Isanbor, Vincent O. Osunkalu, Abimbola A. Sowemimo, Adegoke O. Keshinro, Idowu A. Aneyo, Olawale S. Folarin, Akinbami O. Oladokun, Oluwatosin J. Akinsola, Christianah I. Ayolabi, Tenny O. Egwuatu, Victor A. Owoyomi, Anthony E. Ogbeibu

## Abstract

**Background:** Following the declaration of COVID-19 as a global pandemic and the report of index case in Africa, the number of countries in Africa with confirmed cases of the infection has grown tremendously with disease now being reported in almost all countries on the continent, with the exemption of Lesotho after 75 days. It is therefore necessary to evaluate the disease outcomes among the African countries as the situation unfolds for early identification of best practices for adoption.

**Methods:** In this study, COVID-19 disease outcomes (confirmed cases, deaths and recoveries), testing capacities and disease management approaches among African countries were evaluated. The relationship between COVID-19 infections in African countries and their performance on global resilient indices including the Human Development Index (HDI), performance on Sustainable Development Goals (SDGs) and the Global Risk Index (GRI) were also examined. Data acquired from various standard databases were evaluated over a period of 75 days from the date of reporting the index case.

**Results:** This study has revealed compelling spatial differences in the incidence, deaths and recoveries from COVID-19 among African countries. Egypt, South Africa, Morocco and Algeria were clustered as countries with highest values of COVID-19 disease outcomes on the continent during the 75-day period of observation. The cluster analysis and comparison of countries in terms of percentage recovered cases of confirmed infections revealed that Mauritius, Mauritania, Gambia, Burkina Faso, Madagascar, Togo and Uganda had the highest scores. Comparative analysis of COVID-19 across the world revealed that the parameters were relatively inconsequential in Oceania and Africa continents, while Europe, North America and Asia had significantly higher cases of disease outcomes. For COVID-19 testing capacities, South Africa, Ghana and Egypt are leading in total number of tests carried out. However when the number of tests carried out were related to population number of the countries, Djibouti, Mauritius, Ghana and South Africa are found to be the leading countries. With respect to management of the disease in Africa, all the countries adopted the WHO protocols, personal hygiene, economic palliatives and social distancing measures. Only three countries in Africa (Madagascar, Togo and Burkina Faso) had a state supported initiative to utilise traditional medicines or herbs as alternatives to control COVID-19. Additionally, most of the countries are providing prompt treatment of the patients with a range of drugs especially Hydroxychloroquine, Chloroquine and Chloroquine-Azithromycin combination. The study found that no strong relationship currently exists between the global resilient indicators (HDI, SDG and GRI) and COVID-19 cases across Africa.

**Conclusions:** This study has revealed compelling spatial differences in disease outcomes among African countries and also found testing capacities for COVID-19 to be abysmally low in relation to the population. During the 75 days of observation, African countries have recorded significantly low number of deaths associated with COVID-19 and relatively high recovery rates. Countries in Africa with higher rate of recovery from the disease were found to have adopted strict adherence to some of WHO protocol to contain the disease, isolate all those who test positive to the disease and provide prompt treatment of the patients with a range of drugs especially Hydroxychloroquine, Chloroquine and Chloroquine-Azithromycin combination. The study recommends that the approaches adopted by the African countries which achieved high recovery rates from COVID-19 should be integrated into healthcare management plans for the disease across the continent even as the situation unfolds.

## Introduction

Coronavirus disease 2019 (COVID-19) originating from Wuhan, China, was declared a public health emergency of international concern on 30th January 2020 (Cucinotta and Vanelli, 2020). In Africa, the first COVID-19 confirmed case in Africa was reported in Egypt on the 14^th^ of February 2020 followed by a second reported case in Algeria on the 25^th^ February, 2020 when the Algerian government confirmed that an Italian male, who had entered the country recently had tested positive for COVID-19 (WHO, 2020a, Makoni, 2020). Since the report of index case in Africa, the number of countries in Africa with confirmed cases of the infection has grown tremendously with disease now being reported in almost all countries on the continent, with the exemption of Lesotho where there is currently no official reports of confirmed COVID-19 cases to date (WHO, 2020b).

Testing for Severe Acute Respiratory Syndrome Coronavirus-2 (SARS-CoV-2) in Africa is expected to be very challenging. The Real Time – Polymerase Chain Reaction (RT-PCR) molecular diagnostic technique remains the primary method of diagnosing SARs-CoV2 all over the world and indeed on the African continent. Although RT-PCR remains a very sensitive method of diagnosis, it has been associated with significant cost, averaging 60 dollars per sample (https://allafrica.com/stories). This has been a major setback to extensive testing in Africa. A second challenge is insufficient accredited level 3 and 4 Laboratories for testing and also the low level of manpower required to perform such tests (Nkengasong and Mankoula, 2020). At the beginning of the outbreak, only two referral laboratories – one in South Africa and another in Senegal – were reported to be capable of carrying out the test (WHO, 2020c). The vast majority of national laboratories in Africa lacked the reagents necessary for testing since it is a novel virus. Therefore, testing capabilities needed to be scaled up rapidly in order to give the continent a fighting chance. Although some of the countries have experience responding to communicable diseases like Ebola and Influenza, with 28 countries in the WHO African Region being members of WHO’s Global Influenza Surveillance Network (WHO, 2020c), the testing capabilities for COVID-19 on the continent at the onset of the outbreak was definitely a major source of concern. The huge instant demand for testing reagents and reliable testing kits also put a lot of constraints on supply chains and resulted in production and delivery bottlenecks all around the world. This limited ability to test for the COVID-19 infection was expected to be a major stumbling block for the management of the disease across the continent. It will therefore be necessary to evaluate the testing capacities for COVID-19 infection achieved across the African continent as we enter a phase of quantitative easing of lockdown on the continent, in order to identify best practices, if any, for emulation.

Due to the poor state of health infrastructure, poverty concerns, unfavourable living conditions in the cities, population densities and prevalence of underlying disease conditions like lower respiratory infections, malaria, diarrheal, HIV/AIDS and tuberculosis, there is serious concern that the impact of COVID-19 on the continent will be devastating (The World Bank, 2020; Martinez-Alvarez, 2020). According to UNECA (2020), Africa could see 300,000 deaths from the coronavirus this year even under the best-case scenario. Under the worst-case scenario with no interventions against the virus, Africa could see 3.3 million deaths and 1.2 billion infections. Africa’s largest cities have grown relentlessly for decades to become home to tens of millions of people spread between overcrowded central settlements and offshoots of similarly unplanned sprawl and satellite towns. Countries with higher urban populations are faced with the logistical and communication challenge of informing, monitoring, and possibly isolating a larger pool of at-risk people (Hoogeveen *et al*., 2004). It was feared that the pandemic could be difficult to keep under control in Africa, and could cause huge economic problems if it spreads widely (Gibbs, 2005). Shielding the vulnerable from COVID-19 could involve a mixture of approaches which are traditional and also based on standard medical guidance of early identification of confirmed cases, swift contact tracing with physical isolation, community engagement and focused care, eventually by those who have recovered from the virus (Gilbert *et al*., 2020). These measures will work best when based on local innovations appropriate to particular social contexts and designed with input from those involved. These could build on practices of respect for the elderly and community organising in many African settings, as well as, experiences gained from fighting previous epidemics. It is therefore not surprising that many of the African countries have evolved varied approaches to the management of the diseases with different level of successes. An evaluation of the approaches adopted would merit further investigation.

Africa already suffers from several existing vulnerabilities including: poverty and harsh socio-economic conditions, poor governance, climate change impact, poor resilience, limited access to capital, low level of technology and poor emergency preparedness, among others (Boko *et al*., 2007). In Africa, 33 countries are classified as part of the least developed countries in the world. This classification is based on the evaluation that the countries exhibit the lowest indicators of socioeconomic development, with the lowest human development Index ratings of all countries in the world (United Nations, 1999). Most African countries have fragile health care system which could be easily overwhelmed by a high incidence of the pandemic. Preventive measures that are to be implemented to forestall a catastrophic effect would consequentially have negative impacts on trade and business especially for the informal sector who earn a living daily. Successful response to a global pandemic like COVID-19 will therefore require massive interventions from international agencies like World Health Organization (WHO), The World Bank Group, International Monetary Fund (IMF), African Union (AU), African Development Bank (ADB), African Export-Import Bank (Afrexim bank) and Economic Community of West African States (ECOWAS) to avert a devastating outcome of the disease on the continent. Many of the interventions from the various international organisations must be targeted at preventing unprecedented infection rate, illness and death from COVID-19 and also, to cushion the social disruption and economic consequences of the outbreak in African countries (African Union, 2020). In this regards, the hypothesis therefore, is that the existing vulnerabilities will likely make the COVID-19 pandemic a disaster of epic proportion in Africa. The World Health Organization (WHO) already warned that Africa faces severe danger. It will therefore be necessary to also examine the relationship between COVID-19 infections in African countries and their performance on global resilient indices including the Human Development Index (HDI), performance on Sustainable Development Goals (SDGs) and the Global Risk Index (GRI).

The main objectives of this paper are to provide preliminary evaluation and comparison of the COVID-19 disease outcomes (confirmed cases, deaths and recoveries), testing capacities, disease management approaches and the effects of COVID-19 on global resilient indices across African countries. The paper will also provide the plurality of approaches to manage the disease in Africa and identify lessons learnt.

## Methods

### Study Area

The countries on the continent of Africa were chosen for this study. The categorization of the countries into regions – North Africa, West Africa, East Africa, Central Africa and Southern Africa was also carried out for further evaluation.

### Data Collection Sources

Data on COVID-19 disease outcomes (Confirmed cases of infection, deaths, recoveries), number of tests carried out and population of each country in Africa as at 30^th^ of April 2020 representing 75 days from first index case on the continent in Egypt were sourced from online databases – Worldometer (https://www.worldodometers.info) and COVID-19 dashboard of the Center for Systems Science and Engineering of Johns Hopkins University (https://coronavirus.jhu.edu/map.html). The data were cross checked with data of National Centres for Disease Control for the relevant countries for accuracy.

The case fatality rates and the death per 100,000 of population were computed. The Human Development Index (HDI) data for African countries was sourced from the United Nations Development Programme (UNDP) Human Development Report 2019 (UNDP 2019). The HDI is a composite index composed from main multi-dimensional indices including: life expectancy at birth, expected years of schooling, means years of schooling, and gross national income per capita. The Sustainable Development Goals (SDG) data were sourced from the SDG 2019 Index (Sachs *et al*., 2019). The SDG index is also a composite index consisting of multidimensional indices comprising of scores for all the 17 SDGs. The Global Risk Index (GRI) data was sourced from the INFORM Report 2019 Global Risk Index (INFORM 2019). The index again is a composite index consisting of multi-dimensional indices of several natural, human and socio-economic dimension summarized into: hazard and exposure, vulnerability, and lack of coping capacity.

### Data Analysis

Statistical Analysis of COVID-19 disease outcomes (Confirmed Cases of Infection, Deaths, Recoveries and Tests conducted) across Africa: These data were standardized to percentages using the population figures for the different countries. Multivariate statistics using hierarchical cluster analysis and K-means clustering were performed to categorize countries and group them into clusters on the basis of similarities or disparities using the chosen criteria (Hammer *et al*., 2001; Ogbeibu, 2014).

Spearman’s rank correlation and regression analysis were done to determine relationship between Human Development Index (HDI), Sustainable Development Goals (SDG) index and Global Risk Index (GRI) with COVID-19 disease outcomes data for the study countries. Level of significance is set at 5% (p<0.05).

## Results

### Comparative analysis of COVID-19 disease outcomes among African countries

#### Total Confirmed Cases

Figure 1 shows the number of confirmed cases of COVID-19 in different countries presented in decreasing order, with Egypt, South Africa, Morocco and Algeria having the highest number of confirmed cases when not related to the country’s population. The results of the clustering and classification of the African countries based on total number of confirmed cases are presented in Figures 2 & 3, Table 1. The dendrogram clustering based on Euclidean distance divided the countries into 2 broad groups. Group I comprises of Egypt, South Africa, Morocco and Algeria and Group II comprises of the other countries in Africa. The Group II is divided into 2 further sub-groups which are further divided into 2 smaller groups (Figure 3). Further categorization of the total confirmed cases across Africa using K-means clustering also revealed similar trend though grouping the countries into 3 broad groups (high, medium and low) (Table 1).

**Fig. 1:**
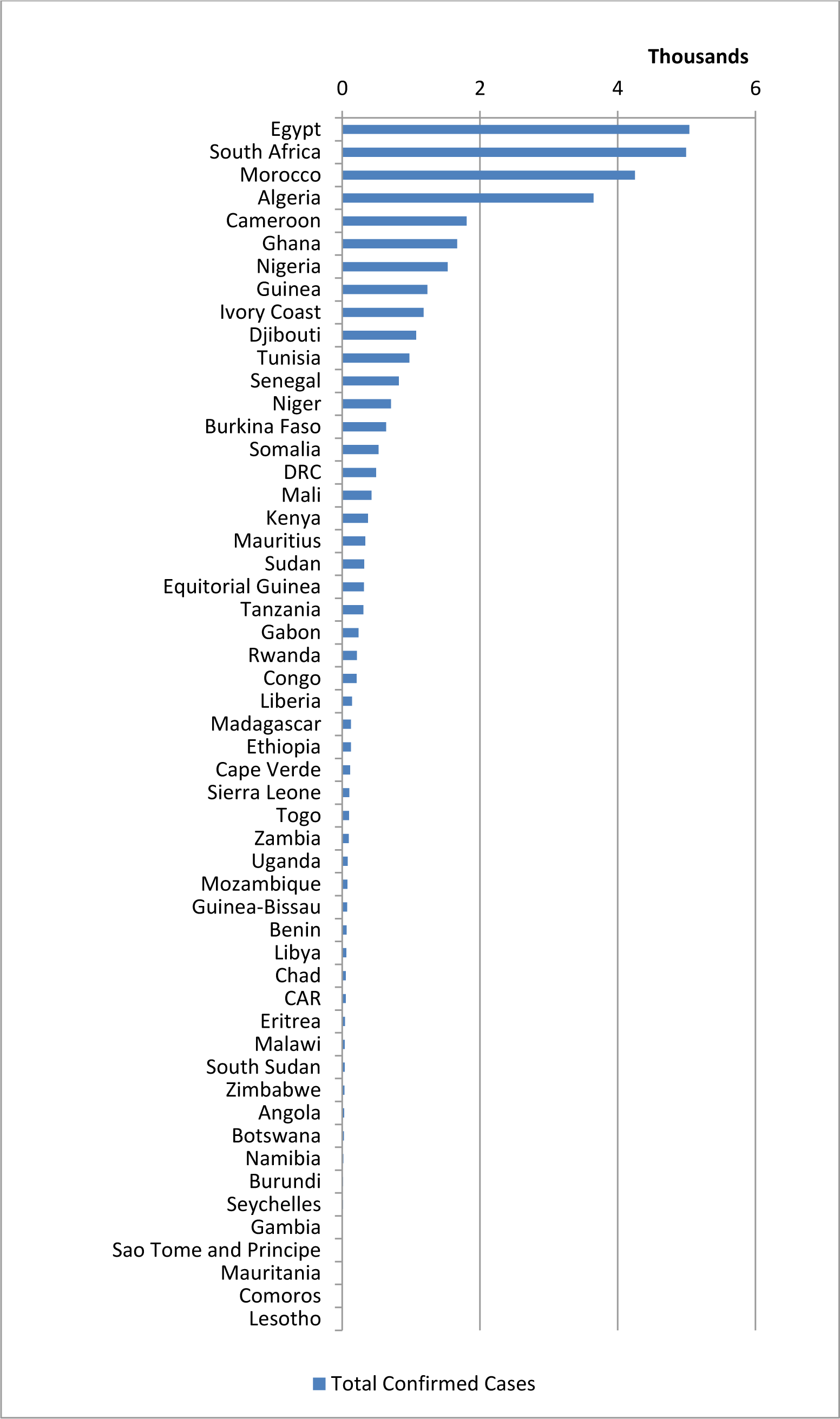
Total confirmed cases associated with COVID-19 among African countries.

**Fig. 2:**
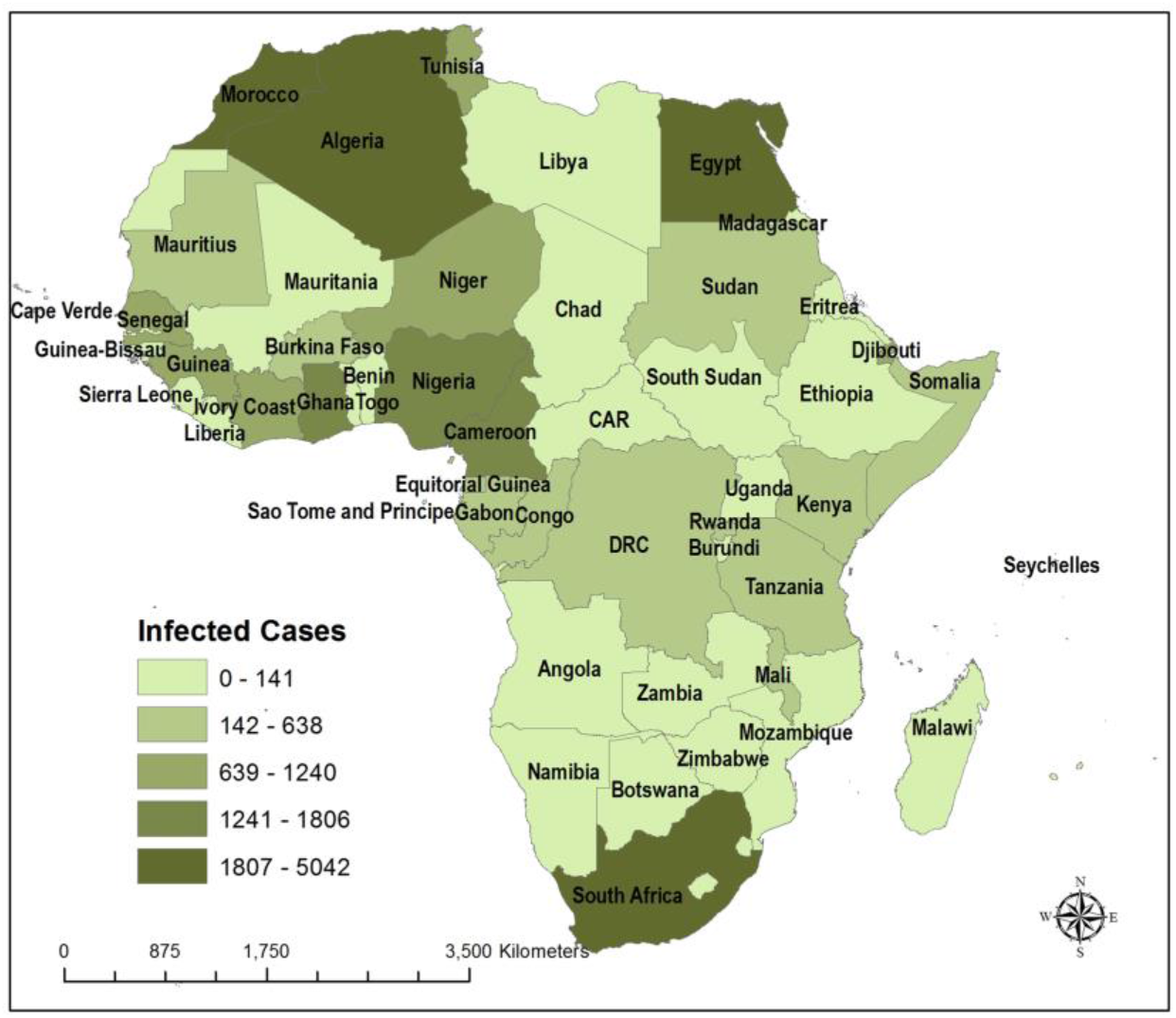
COVID-19 confirmed cases of infection across Africa (as at 30 April 2020)

**Fig. 3:**
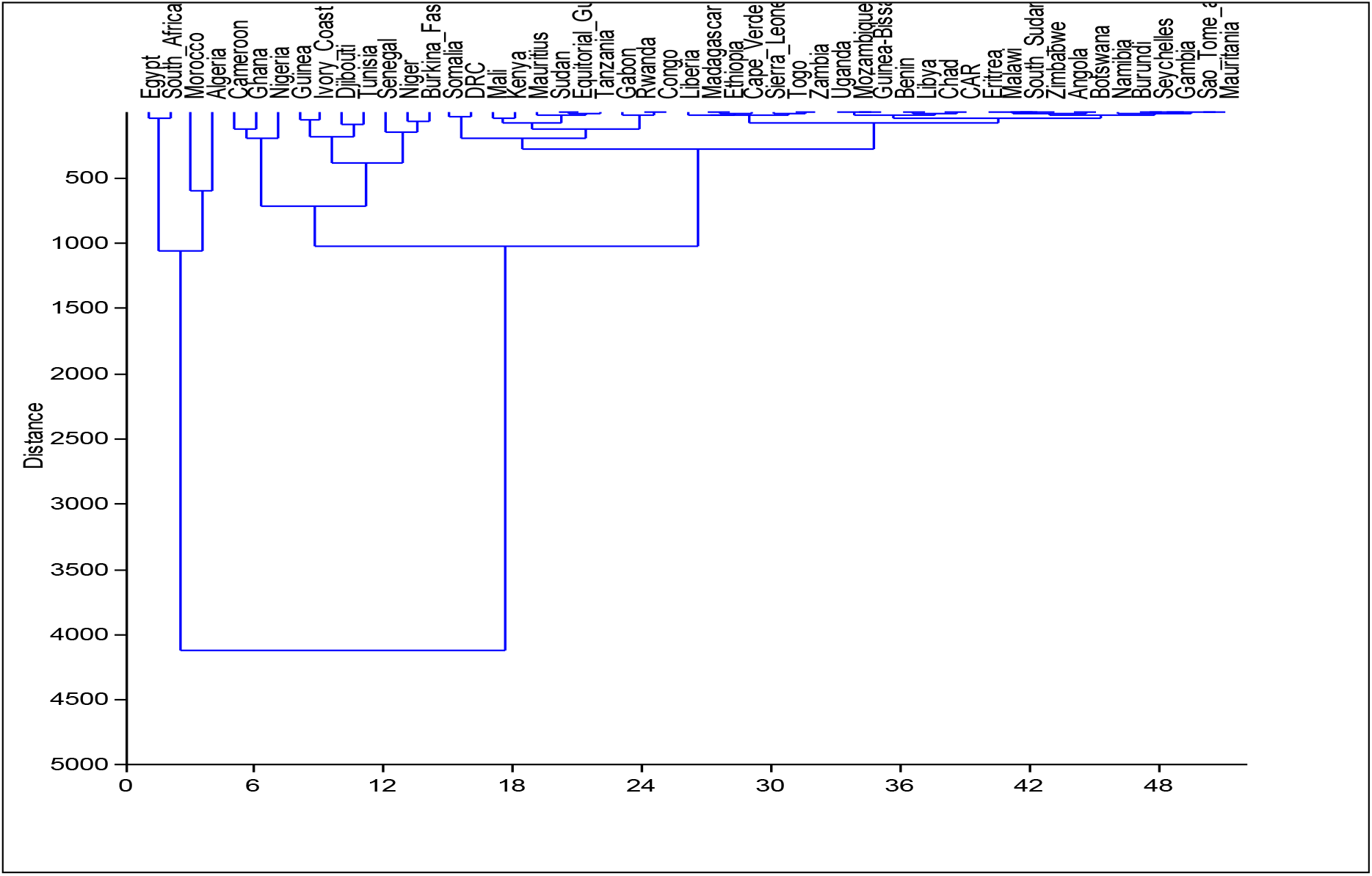
Hierarchical cluster analysis of similarities among African countries based on total confirmed cases of COVID-19 using Euclidean distance.

**Table 1:** Classification of confirmed cases using K-means clustering.

| **HIGH (2000-5500)** | **MEDIUM (700-1999)** | **LOW (<700)** |  |
| --- | --- | --- | --- |
| Egypt | Cameroon | Burkina_Faso | Uganda |
| South_Africa | Ghana | Somalia | Mozambique |
| Morocco | Nigeria | DRC | Guinea-Bissau |
| Algeria | Guinea | Mali | Benin |
|  | Ivory_Coast | Kenya | Libya |
|  | Djibouti | Mauritius | Chad |
|  | Tunisia | Sudan | CAR |
|  | Senegal | Equitorial_Guinea | Eritrea |
|  | Niger | Tanzania | Malawi |
|  |  | Gabon | South_Sudan |
|  |  | Rwanda | Zimbabwe |
|  |  | Congo | Angola |
|  |  | Liberia | Botswana |
|  |  | Madagascar | Namibia |
|  |  | Ethiopia | Burundi |
|  |  | Cabo_Verde | Seychelles |
|  |  | Sierra_Leone | Gambia |
|  |  | Togo | Sao_Tome_and_Principe |
|  |  | Zambia | Mauritania |

#### Total Deaths

The result of total deaths associated to COVID-19 among African countries is present in Figure 4. Algeria, Egypt and Morocco have the highest number of death at 437, 359 and 165 respectively. Other countries had less than 100 COVID-19 related deaths during the period of study. The results of the clustering and classification of the African countries based on total death numbers are presented in Figure 5 and Table 2. The dendrogram clustering based on Euclidean distance divided the countries into 2 broad groups. Group I comprises Egypt, South Africa, Morocco and Algeria and Group II comprises the other countries in Africa. The Group II is divided into 2 further sub-groups – Cameroon and others, within the others, there are several other subdivisions (Figure 5). Further categorization of the countries based on the total death cases of COVID-19 across Africa using K-means clustering revealed three broad groups – High, Medium and Low. Egypt and Algeria are in the high group; South Africa, Morocco and Cameroon represented in medium group and other remaining countries are in the low group (Table 2).

**Fig. 4:**
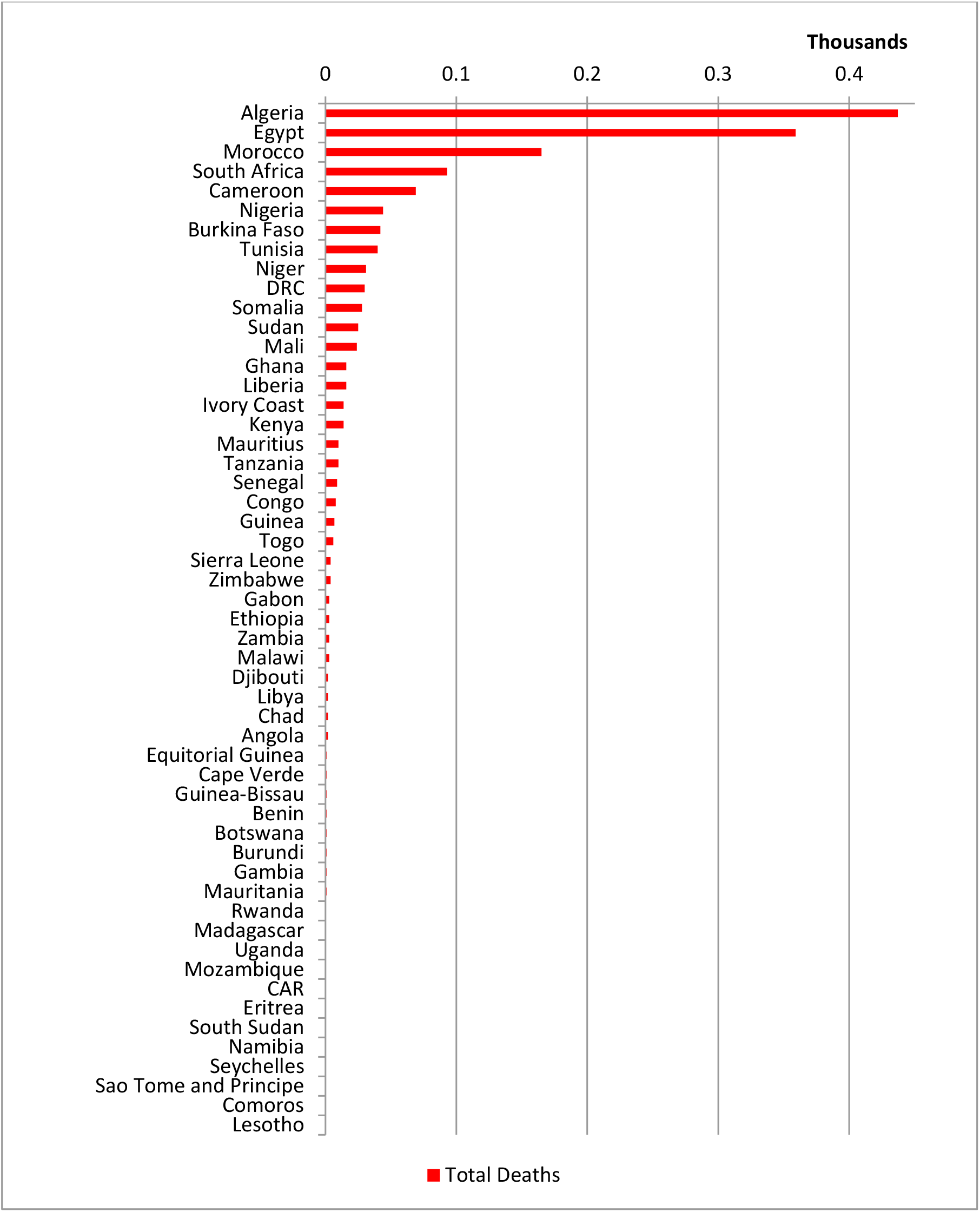
Total of deaths associated with COVID-19 among African countries.

**Fig. 5:**
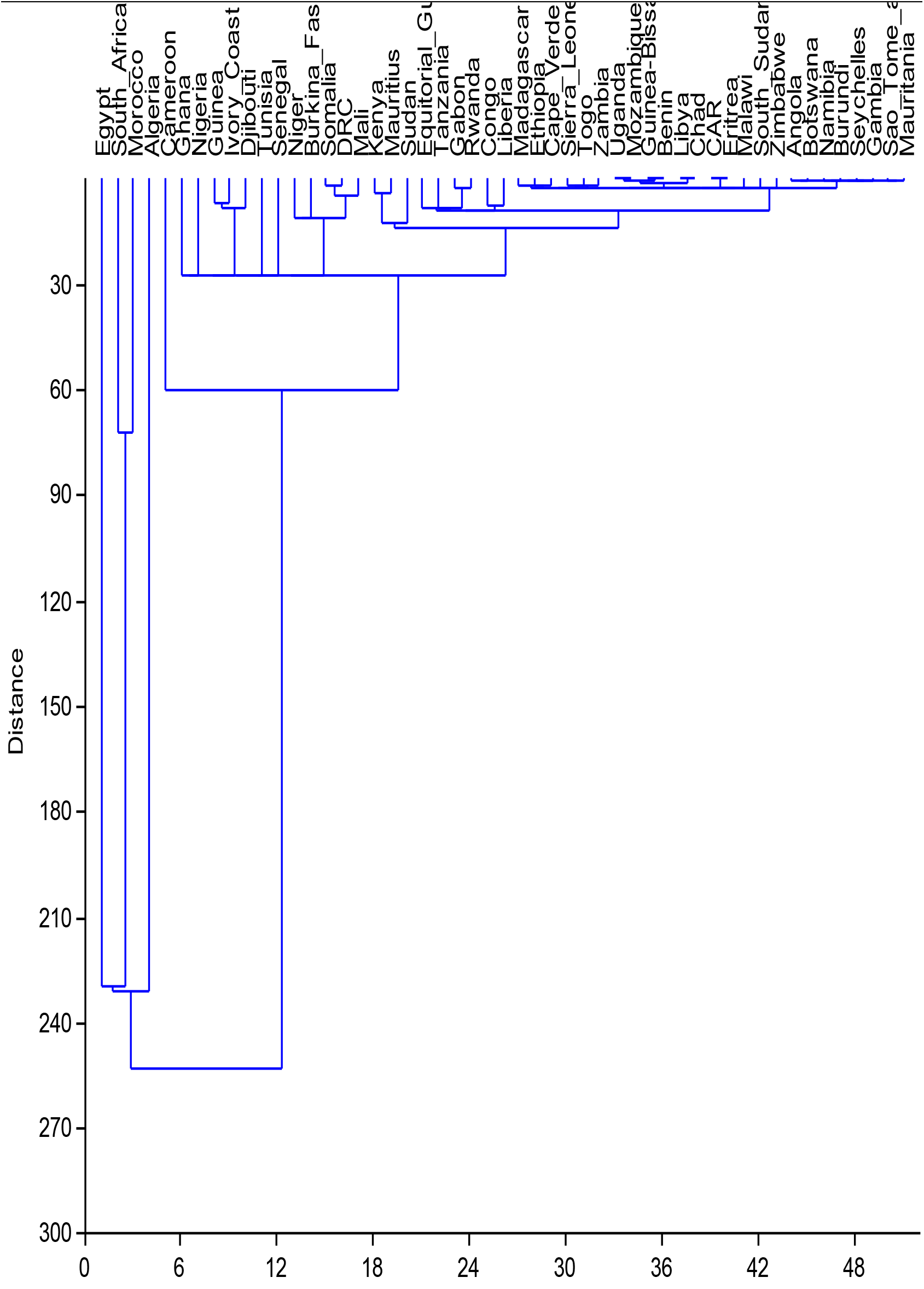
Hierarchical cluster analysis of similarities among African countries based on total death cases of COVID-19 using Euclidean distance.

**Table 2:** Classification of COVID-19 deaths number using K-means clustering.

| **HIGH (>300)** | **MEDIUM (50-100)** | **LOW (<50)** |  |
| --- | --- | --- | --- |
| Egypt | South Africa | Ghana | Cabo_Verde |
| Algeria | Morocco | Nigeria | Sierra_Leone |
|  | Cameroon | Guinea | Togo |
|  |  | Ivory_Coast | Zambia |
|  |  | Djibouti | Uganda |
|  |  | Tunisia | Mozambique |
|  |  | Senegal | Guinea-Bissau |
|  |  | Niger | Benin |
|  |  | Burkina_Fa | Libya |
|  |  | Somalia | Chad |
|  |  | DRC | CAR |
|  |  | Mali | Eritrea |
|  |  | Kenya | Malawi |
|  |  | Mauritius | South_Sudan |
|  |  | Sudan | Zimbabwe |
|  |  | Equitorial_ | Angola |
|  |  | Tanzania | Botswana |
|  |  | Gabon | Namibia |
|  |  | Rwanda | Burundi |
|  |  | Congo | Seychelles |
|  |  | Liberia | Gambia |
|  |  | Madagascar | Sao_Tome_and_Principe |
|  |  | Ethiopia | Mauritania |

#### Recovered Cases

Figure 6 depicts the spatial variations in total recovered cases in relation to total confirmed cases and deaths among African countries. South Africa, Algeria and Egypt have the highest number of total recovery at 2,073, 1,651 and 1,304 respectively. The comparison of countries in terms of percentage recovered cases of confirmed infections however revealed that Mauritius, followed by Mauritania and Gambia had the highest scores (Figure 7). Only 13 countries out of 51 had greater than 50% recovered cases. About 75% of the countries investigated had less than 50% of recovered cases. The results of the clustering and classification of the African countries based on percentage recovered cases are presented in Figure 8 and Table 3. The dendrogram clustering based on Euclidean distance divided the countries into 2 broad groups. Group I comprises of the following countries which are subdivided as follows: Burkina Faso and Gambia, Mauritius and Mauritania, Madagascar and Togo, Uganda. All other countries are in Group 2 which is further divided into two broad divisions with several other subdivisions (Figure 8). Further categorization of the countries based on the percentage recovered cases of COVID-19 across Africa using K-means clustering revealed three broad groups – High, Medium and Low. Similarly, Mauritius, Mauritania, Gambia, Burkina Faso, Uganda, Madagascar and Togo are in the high group, other countries are in medium and low groups (Table 3).

**Table 3:** Classification of percentage recovered cases using K-means clustering.

| HIGH (60-100%) | MEDIUM (20-59%) |  | LOW (0-19%) |
| --- | --- | --- | --- |
| Mauritius | Niger | Senegal | Zimbabwe |
| Mauritania | Seychelles | Kenya | Morocco |
| Gambia | Benin | Liberia | Nigeria |
| Burkina_Faso | Cameroon | Libya | Mozambique |
| Uganda | Namibia | Mali | Tanzania |
| Madagascar | Sao_Tome_and_Principe | Tunisia | Malawi |
| Togo | Eritrea | Egypt | DRC |
|  | Djibouti |  | Sierra_Leone |
|  | Algeria |  | Ghana |
|  | Rwanda |  | Sudan |
|  | Ivory_Coast |  | Congo |
|  | Zambia |  | Somalia |
|  | South_Africa |  | Equitorial_Guinea |
|  | Ethiopia |  | Cabo_Verde |
|  | Chad |  | South_Sudan |
|  | Burundi |  | Botswana |

**Fig. 6:**
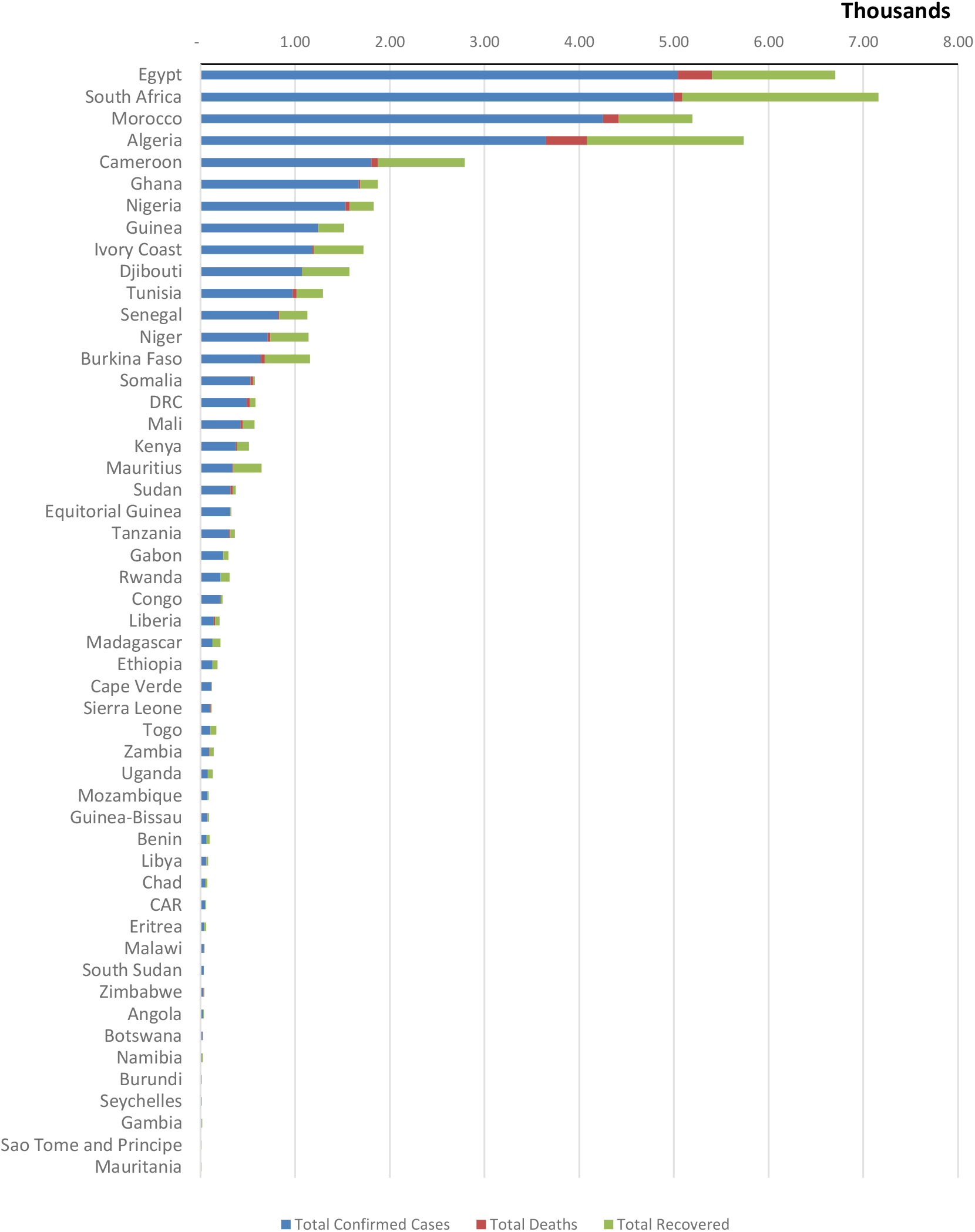
Spatial variation in total confirmed cases, deaths and recovered COVID-19 cases among African countries.

**Fig. 7:**
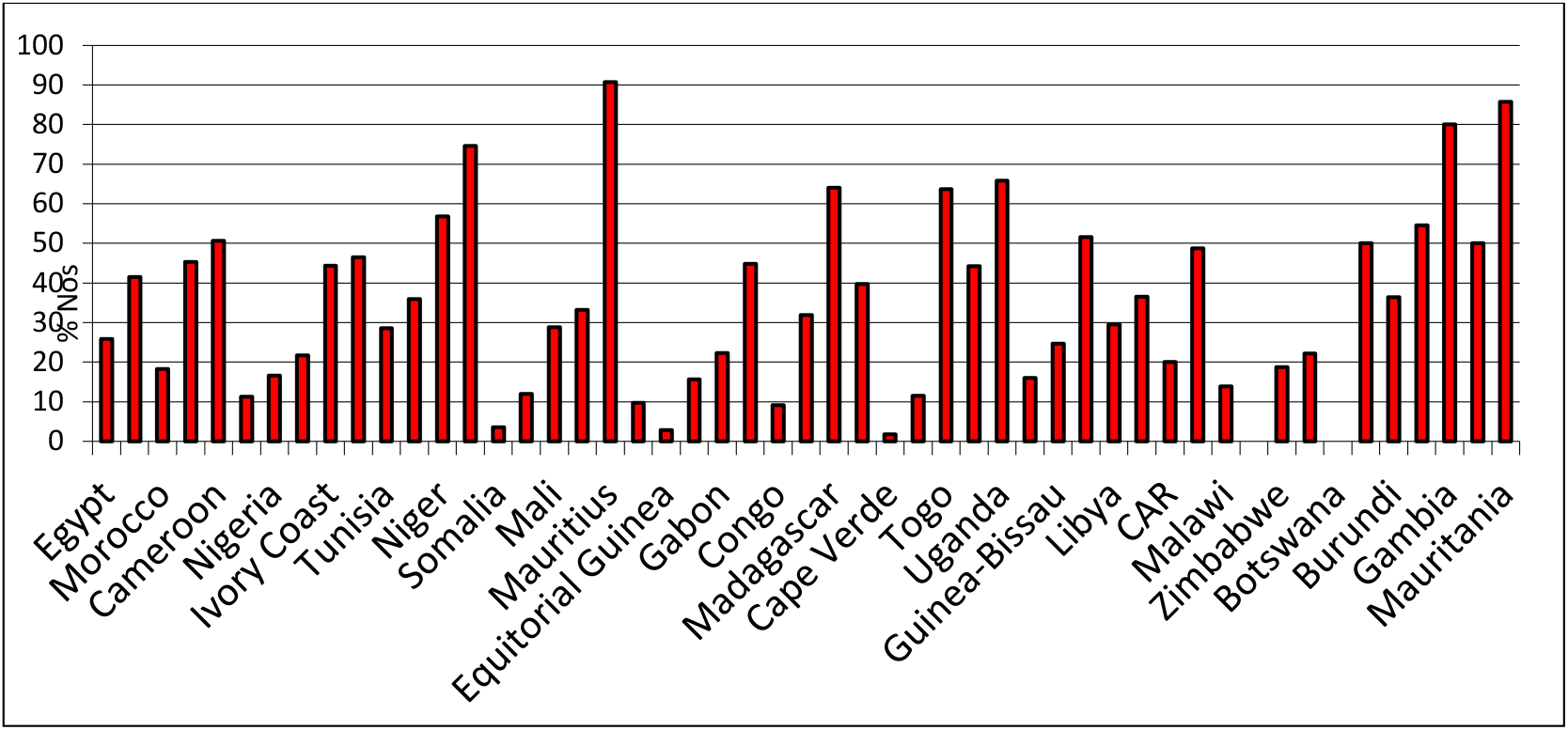
Percentage recovered cases in different countries in Africa in relation to total confirmed cases.

**Fig. 8:**
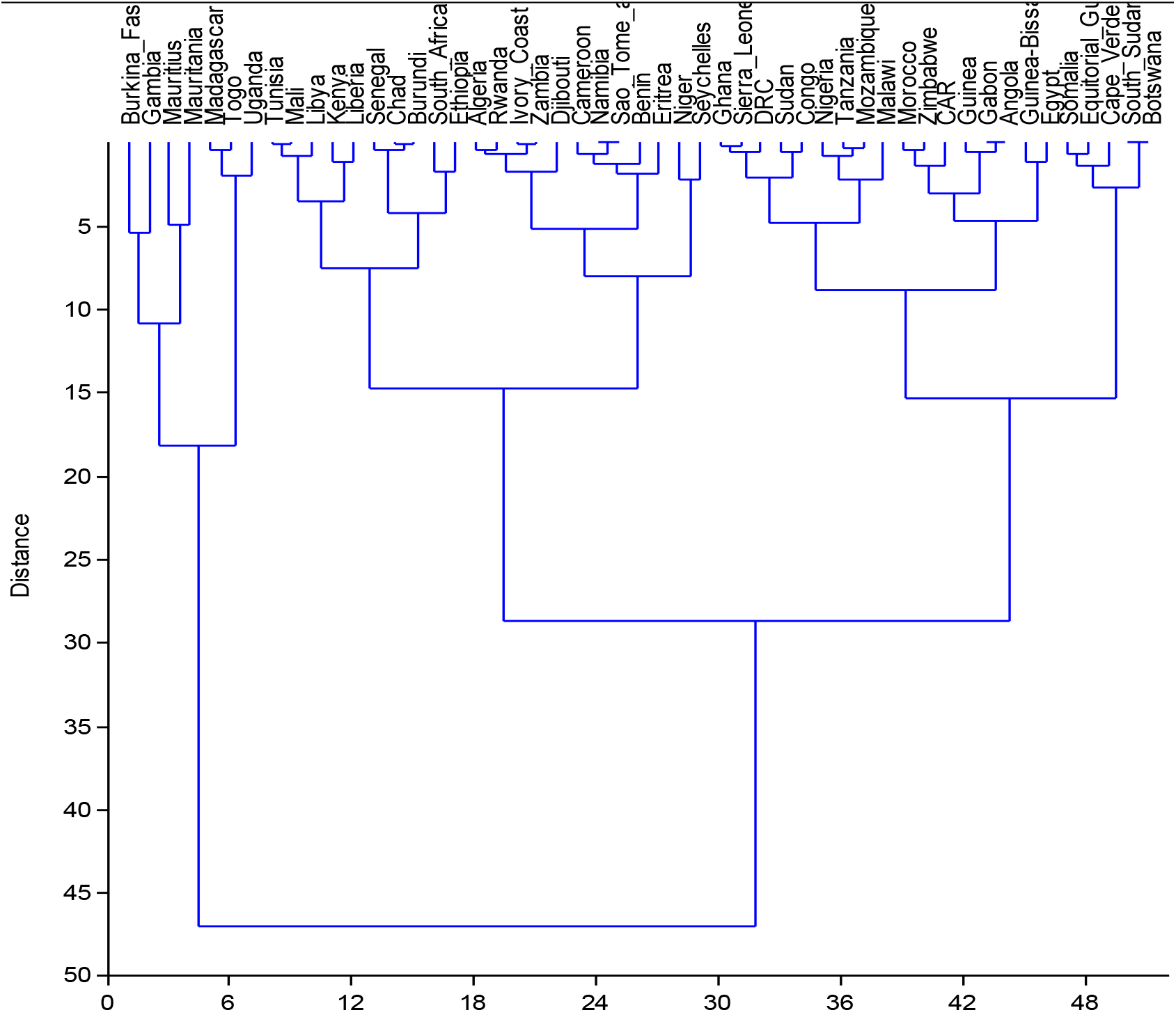
Hierarchical cluster analysis of similarities among African countries based on total recovered cases of COVID-19 using Euclidean distance.

### Comparative analysis of COVID-19 disease outcomes across African regions

The results of the COVID-19 disease outcomes across the African regions are provided in Figure 9. The results depict that North Africa has the highest number of confirmed cases, and deaths followed by West Africa, Southern Africa, East Africa and Central Africa (least affected).

**Fig. 9:**
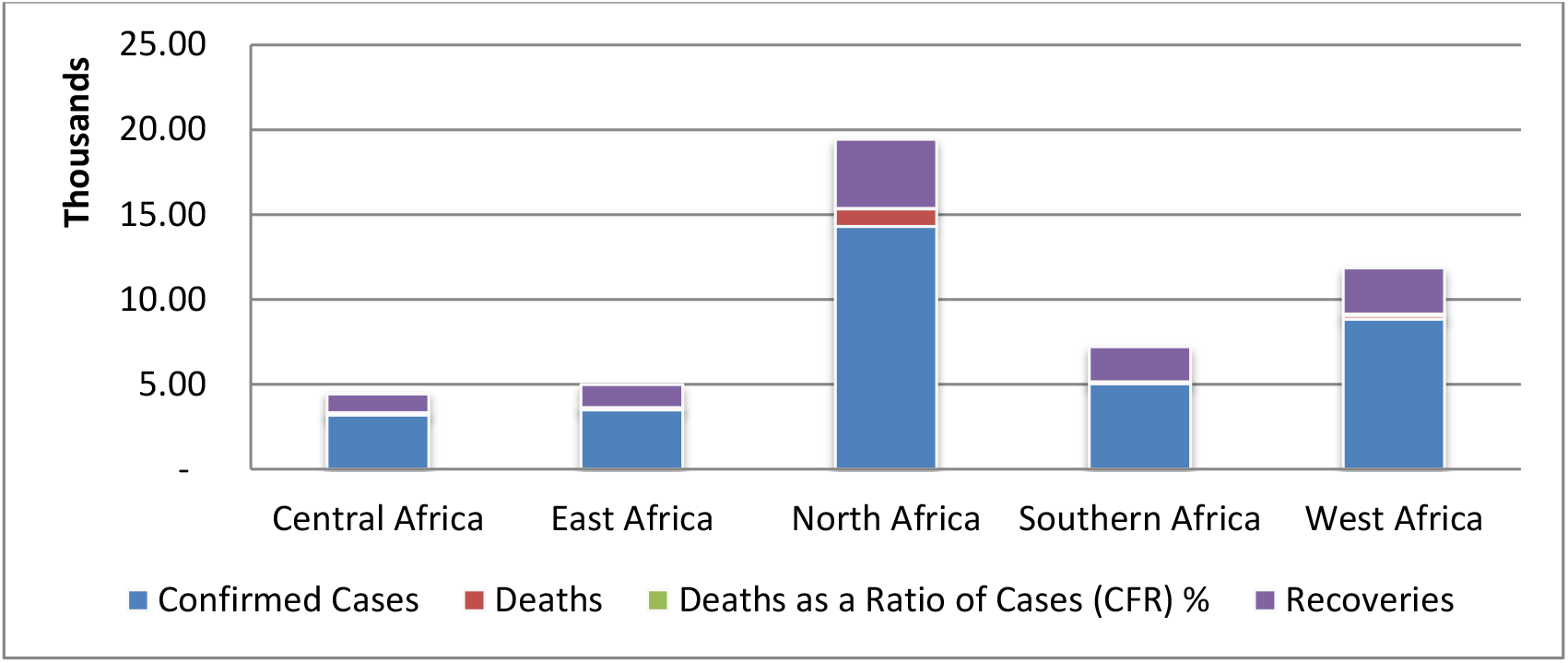
Variation in total confirmed cases, deaths and recovered COVID-19 cases across African regions.

### Comparison of COVID-19 disease outcomes across continents

A comparative analysis of covid-19 disease outcomes (total confirmed cases, total deaths and total recovered) across continents (Fig. 10) revealed that these evaluation parameters were relatively inconsequential in Oceania and Africa, while Europe, North America and Asia had significantly higher cases of disease outcomes.

**Fig. 10:**
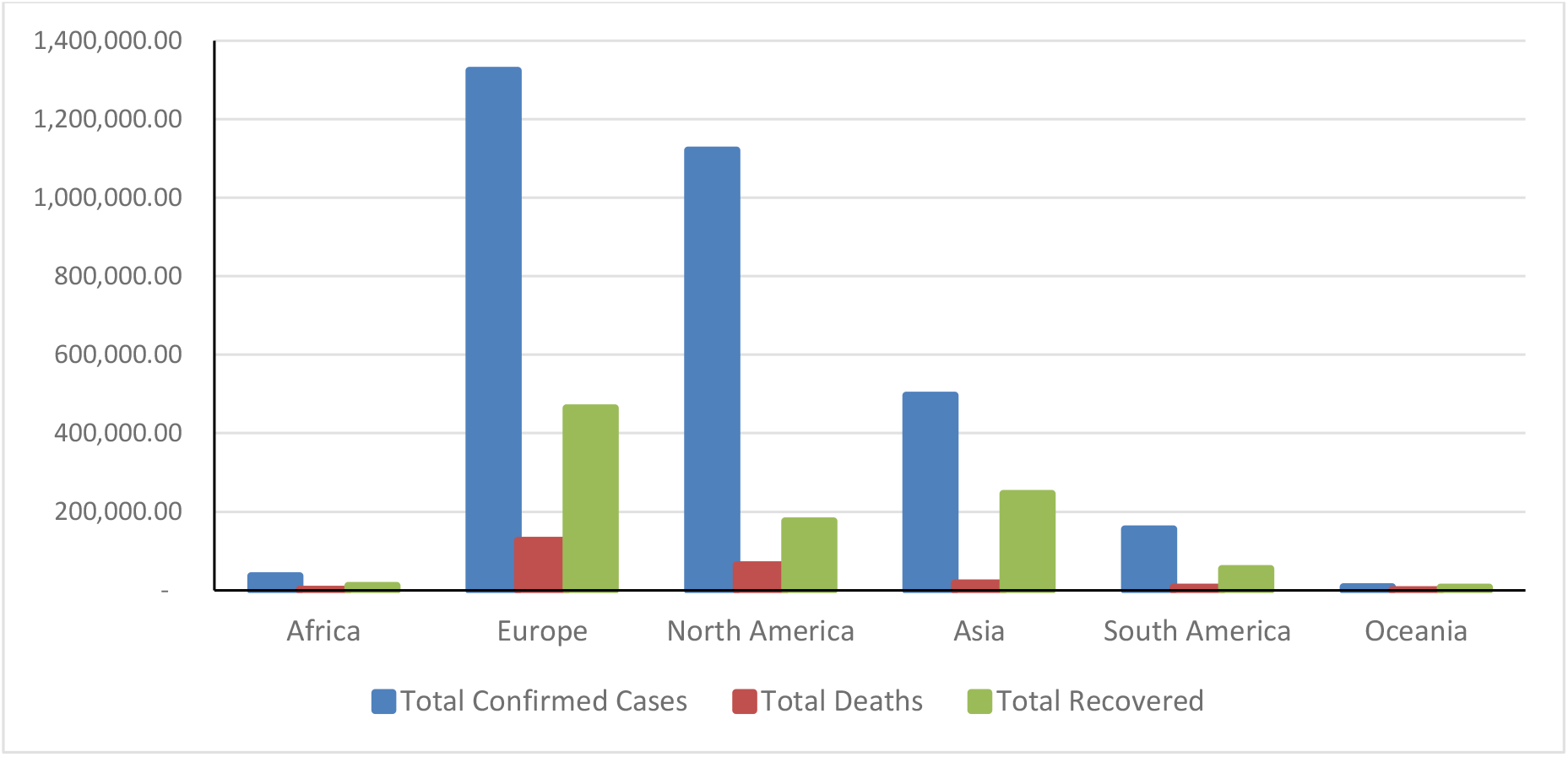
Comparisons of COVID-19 disease outcomes across continents.

### Tests Capacity for COVID-19 among African countries

The results of the COVID-19 testing capacities across African countries are presented in Table 4 and Figures 11 and 12. The results showed in terms of total number of tests carried out, South Africa, Ghana and Egypt are leading in COVID-19 test with 185,497, 100,622 and 90,000 respectively (Fig. 11). However when the number of tests carried out were related to population number of the countries, Djibouti, Mauritius, Ghana and South Africa are found to be leading with 1530, 1136, 324 and 313 per 100,000 of the population respectively. A second group of the countries including Botswana, Tunisia, Seychelles, Cabo Verde and Guinea have testing capacity of 256, 182, 158, 142 and 133 per 100,000 of the population respectively. All other countries have testing capacities below 100 per 100,000 of their population (Fig.12). The results of the clustering and classification of the African countries based on percentage tests per population of the country are presented in Figure 13 and Table 4. The dendrogram clustering based on Euclidean distance put all the countries in Africa in one group which is further divided into 3 subdivisions (Figure 13). Further categorization of the countries based on the percentage COVID-19 tests carried out per population of each countries using K-means clustering revealed three broad groups – High comprising only Mauritius, Medium comprising Ghana, South Africa, Botswana, Tunisia and Cabo Verde and Low which comprises the remaining other countries (Table 3).

**Table 4:**
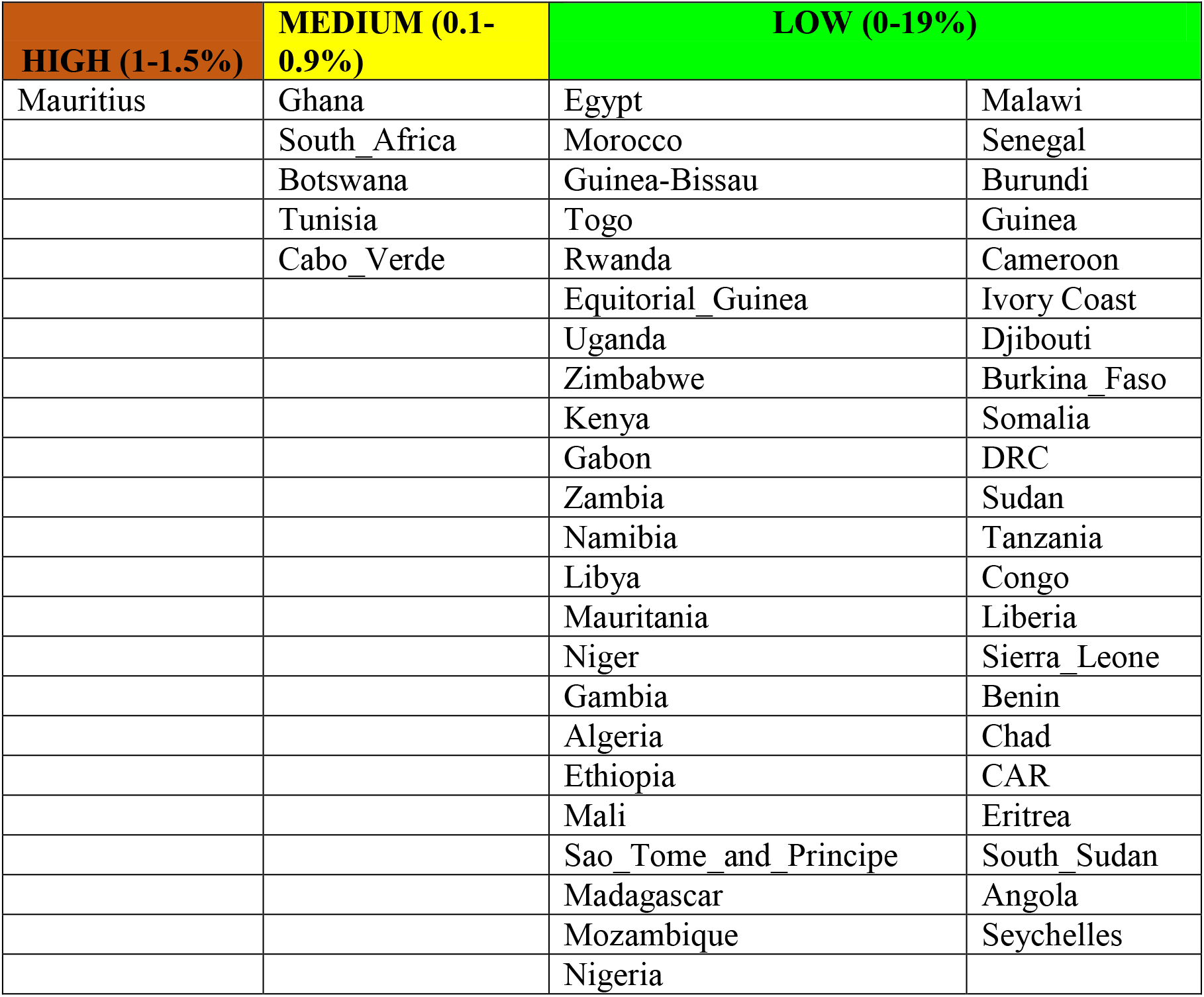
Classification of percentage of population tested using K-means clustering.

**Fig. 11:**
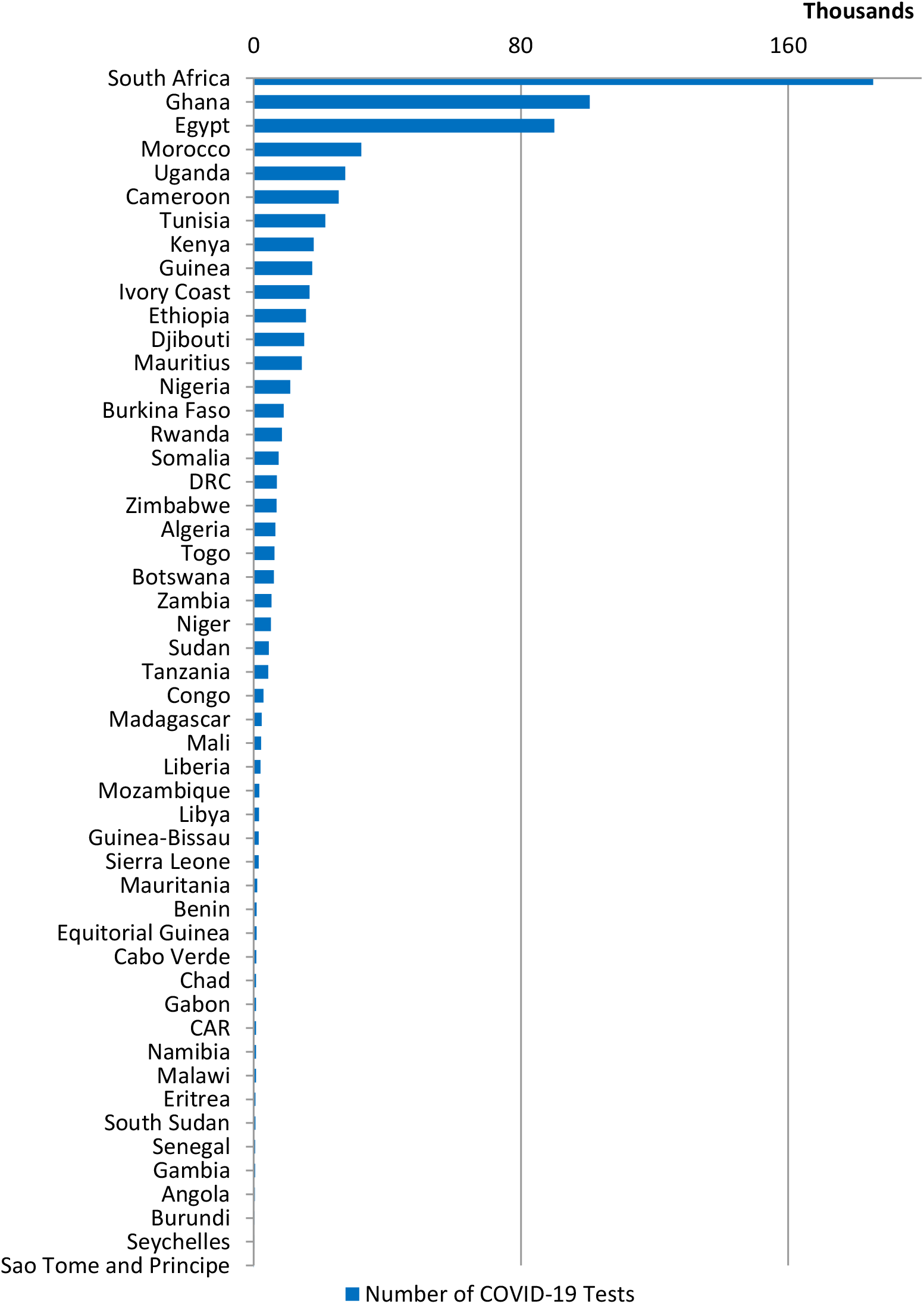
Total number of COVID-19 tests carried out among African countries.

**Fig. 12:**
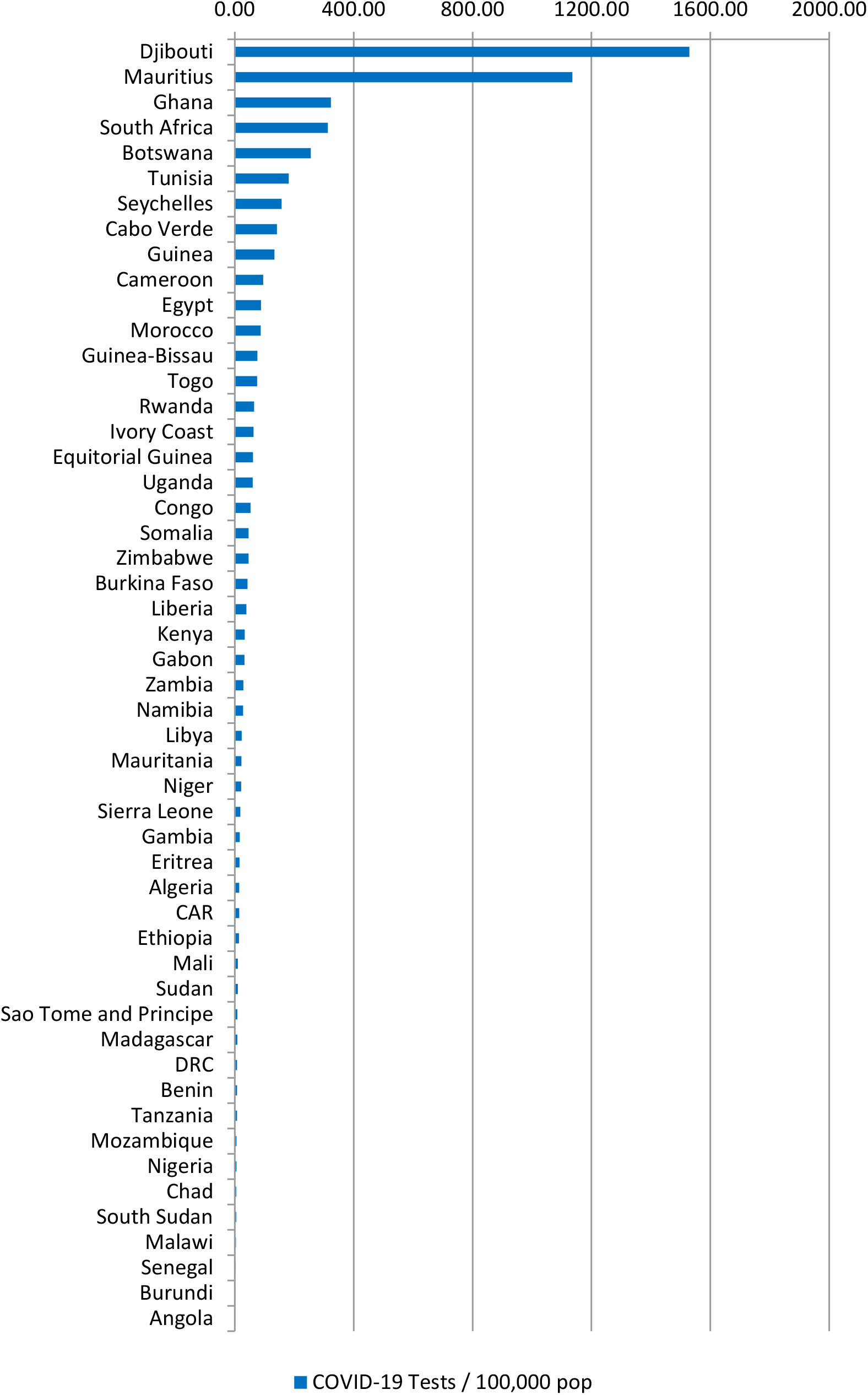
COVID-19 tests per population of African countries.

**Fig. 13:**
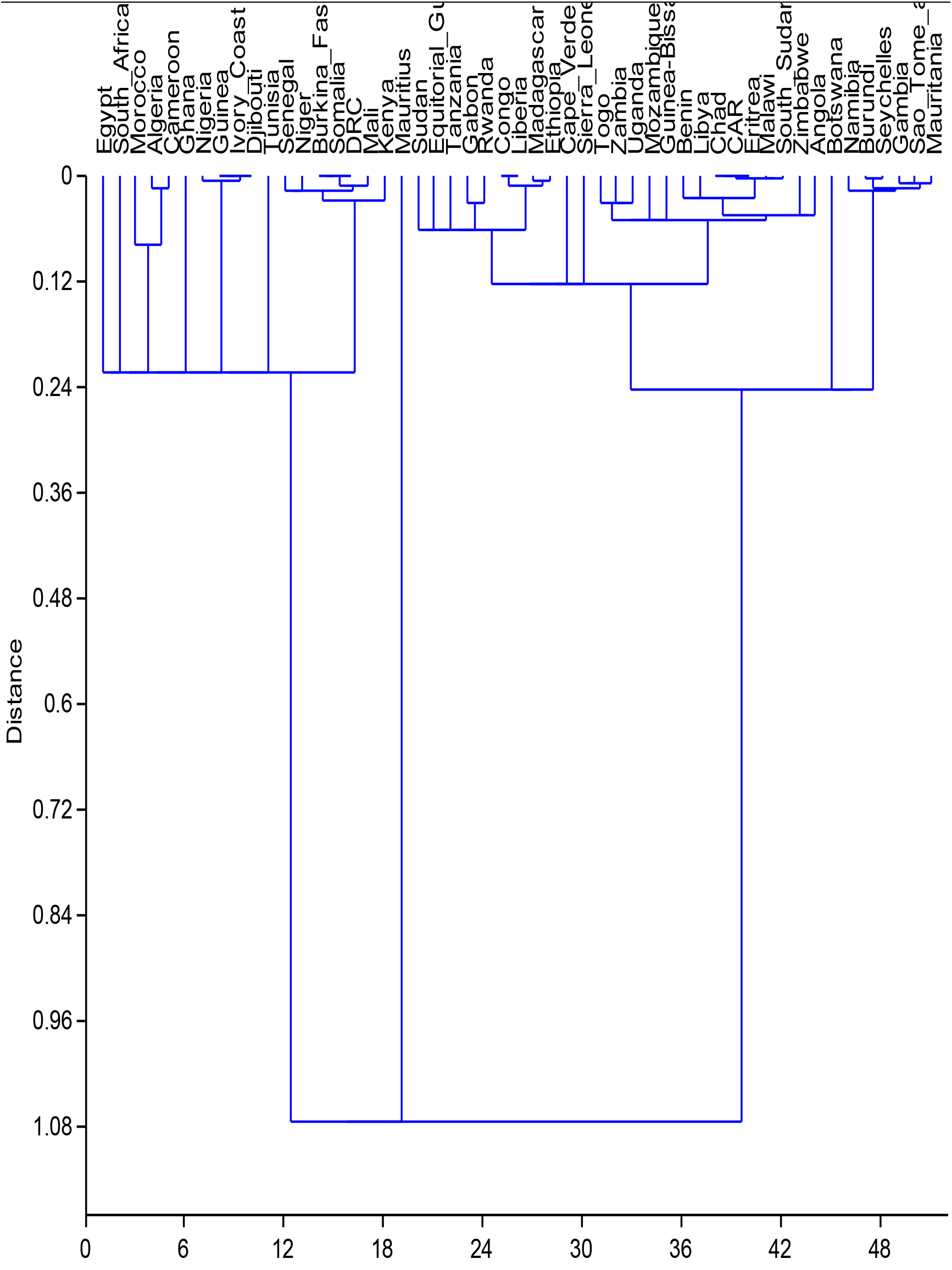
Hierarchical cluster analysis of similarities among African countries based on percentage of population tested for COVID-19 using Euclidean distance.

### Healthcare approaches and treatment of COVID-19 in Africa

A survey of the different approaches adopted by various countries on the African continent revealed a plurality of approaches and treatment for management of COVID-19. The approaches ranges from adopting WHO protocol and guidance (test, isolate & contact tracing), support initiatives on use traditional medicines / herbs, compulsory use of face masks in public places, establishment of COVID-19 of isolation centres, personal hygiene, instituting economic palliative measures, social distancing, international border closure and national lockdown (Table 5).

**Table 5:** Approaches adopted by African countries for management of COVID-19.

| **Countries** | **\*AWP** | **\*TMH** | **\*FMPP** | **\*CIS** | **\*PHM** | **\*EPM** | **\*SD** | **\*IBC** | **\*NLD** | **Innovative Approaches / Remarks** |
| --- | --- | --- | --- | --- | --- | --- | --- | --- | --- | --- |
| *Group I - Countries | X | - | NC | X | X | X | X | X | X | Rampant treatment of symptoms with Chloroquine, Hydroxychloroquine, antibiotics etc |
| *Group II - Countries | X | - | C | X | X | X | X | X | X | Rampant treatment of symptoms with Chloroquine, Hydroxychloroquine, antibiotics etc |
| Ghana | X | - | C | X | X | X | X | X | X | Real-time COVID-19 tracker. Rapid testing kit. Zipline - delivery drones for COVID-19 test in rural areas |
| Kenya | X | - | C | X | X | X | X | X | X | Development of Rapid Test Kits |
| Lesotho | X | - | NC |  | X | X | X | X | X | No reported COVID-19 Case |
| Madagascar | X | X | C | X | X | X | X | X | X | Madagascar launched herbal medicine (COVID-organics) against COVID-19 as prophylaxis. |
| Malawi | X | - | NC |  | X | X | X | - | - | No Lockdown |
| Sao Tome and Principe | X | - | NC |  | X | X | X | - | - | No Lockdown |
| Senegal | X | - | NC | X | X | X | X | X | - | Development of Rapid Test Kits Costing 1 dollar. 3D printing |

| Countries | *AWP | *TMH | *FMPP | *CIS | *PHM | *EPM | *SD | *IBC | *NLD | Innovative Approaches / Remarks |
| --- | --- | --- | --- | --- | --- | --- | --- | --- | --- | --- |
|  |  |  |  |  |  |  |  |  |  | manufacturing of ventilators.<br>Chloroquine<br>Hydroxychloroquine<br>No lockdown,<br>Curfew only – 8pm to 6am. |
| Mauritius | X | - | C | X | X | X | X | X | X | Serum Plasma Therapy<br>Hydroxychloroquine |
| Uganda | X | - | C | X | X | X | X | X | X | Hydroxychloroquine |
| Togo | X | X | C | X | X | X | X | X | X | Chloroquine<br>Traditional medicine use encourages |
| Mauritania | X | - | NC | X | X | X | X | - | - | Strong response team to test, isolate, trace and treatment.<br>No lockdown only night curfew |
| Gambia | X | - | NC | X | X | X | X | X | X | Introduced price control.<br>Hydroxychloroquine |
| Burkina Faso | X | X | C | X | X | X | X | X | X | -Chloroquine<br>-Chloroquine-Azithromycin<br>-Herbal drug - Apivirine (Api-COVID-19) |
| South Africa | X | - | C | X | X | X | X | X | X | COVID-19 test kits that give results in an hour. |

Group I - Algeria, Angola, Botswana, Burundi, Cabo Verde, Central African Republic (CAR), Chad, Comoros, Congo, Djibouti, Eritrea, Eswatini, Gambia, Guinea-Bissau & Libya
Group II - Benin, Burkina Faso, Cameroon, Congo Democratic Republic, Cote d'Ivoire, Egypt, Equatorial Guinea, Ethiopia, Gabon, Guinea, Kenya, Liberia, Morocco, Mozambique, Namibia, Nigeria, Rwanda, Sierra Leone, Zambia
Legends: C - Compulsory use of face mask in public, NC - Face mask in use but not compulsory, AWP - Adopted WHO Protocol, TMH - Traditional Medicines / Herbs, FMPP - Face masks in public places, CIS - COVID-19 Isolation Centres, PHM - Personal Hygiene Measures, EPM - Economic Palliative Measures, SD - Social Distancing, IBC - International Border Closure, NLD - National Lockdown

All countries considered in Africa adopted WHO protocols, personal hygiene, economic palliatives and social distancing measures. Only three countries in Africa (Madagascar, Togo and Burkina Faso) had a state supported initiative to utilize traditional medicines or herbs as alternatives to control COVID-19. A lot of the countries have introduced the use of face masks in public places. Almost all countries in Africa enforced the closure of international borders except Malawi, Sao Tome and Principe and Mauritania. Similarly most countries adopted lockdown measures except Malawi, Sao Tome and Principe, Mauritania and Senegal which only implemented dusk to dawn curfew. Most countries in Africa also engaged in compulsory isolation and treatment of people who tested positive for COVID-19 with mainly hydroxychloroquine, Chloroquine and Chloroquine-Azithromycin combination. Madagascar was reported to have used a herbal tonic, COVID-Organics as a prophylaxis for its population while Burkina Faso put into clinical trial a herbal drug, Apivirine (Api-COVID-19) which is purported to have antiviral properties. Mauritius was also reported to have successfully used the Serum Plasma Therapy for people with severe cases (Table 5).

### Effects of COVID-19 disease outcomes on global resilient indices (GRI) in Africa

In order to reduce impact of COVID-19 on some of the poor global resilient index countries in Africa, a number of interventions and initiatives were introduced at the onset of the outbreak to offset the potential impact. Some of the initiatives have been compiled in table 6.

**Table 6:**
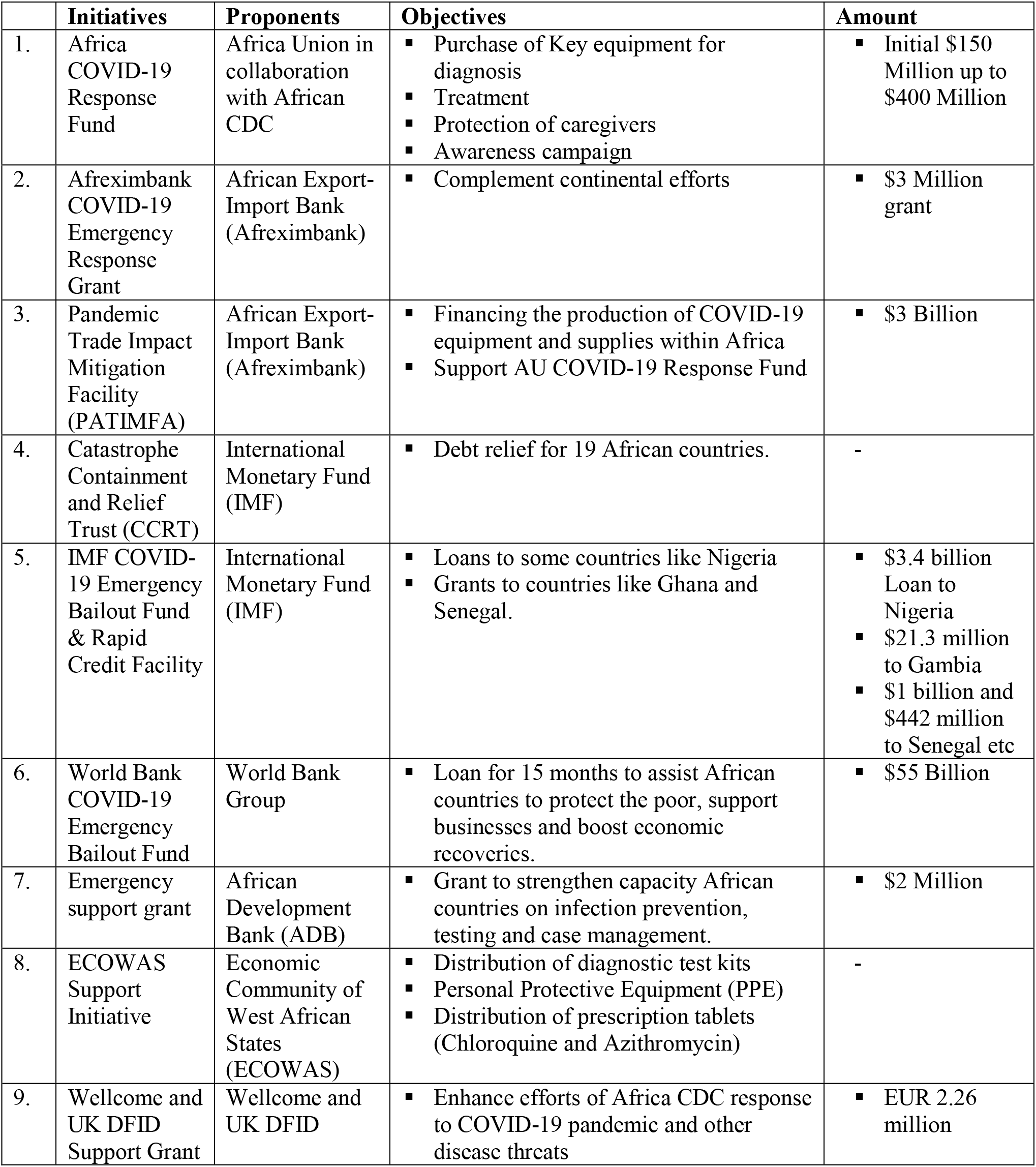
Multilateral and bilateral agencies interventions to COVID-19 pandemic in Africa.

|  | **Initiatives** | **Proponents** | **Objectives** | **Amount** |
| --- | --- | --- | --- | --- |
| 1. | Africa COVID-19 Response Fund | Africa Union in collaboration with African CDC | ▪ Purchase of Key equipment for diagnosis<br>▪ Treatment<br>▪ Protection of caregivers<br>▪ Awareness campaign | ▪ Initial \$150 Million up to \$400 Million |
| 2. | Afreximbank COVID-19 Emergency Response Grant | African Export-Import Bank (Afreximbank) | ▪ Complement continental efforts | ▪ \$3 Million grant |
| 3. | Pandemic Trade Impact Mitigation Facility (PATIMFA) | African Export-Import Bank (Afreximbank) | ▪ Financing the production of COVID-19 equipment and supplies within Africa<br>▪ Support AU COVID-19 Response Fund | ▪ \$3 Billion |
| 4. | Catastrophe Containment and Relief Trust (CCRT) | International Monetary Fund (IMF) | ▪ Debt relief for 19 African countries. | - |
| 5. | IMF COVID-19 Emergency Bailout Fund & Rapid Credit Facility | International Monetary Fund (IMF) | ▪ Loans to some countries like Nigeria<br>▪ Grants to countries like Ghana and Senegal. | ▪ \$3.4 billion Loan to Nigeria<br>▪ \$21.3 million to Gambia<br>▪ \$1 billion and \$442 million to Senegal etc |
| 6. | World Bank COVID-19 Emergency Bailout Fund | World Bank Group | ▪ Loan for 15 months to assist African countries to protect the poor, support businesses and boost economic recoveries. | ▪ \$55 Billion |
| 7. | Emergency support grant | African Development Bank (ADB) | ▪ Grant to strengthen capacity African countries on infection prevention, testing and case management. | ▪ \$2 Million |
| 8. | ECOWAS Support Initiative | Economic Community of West African States (ECOWAS) | ▪ Distribution of diagnostic test kits<br>▪ Personal Protective Equipment (PPE)<br>▪ Distribution of prescription tablets (Chloroquine and Azithromycin) | - |
| 9. | Wellcome and UK DFID Support Grant | Wellcome and UK DFID | ▪ Enhance efforts of Africa CDC response to COVID-19 pandemic and other disease threats | ▪ EUR 2.26 million |

Twenty (20) of the 51 African countries included in this analysis fall under the low human development category with HDI of 0.507 or less. 19 countries fall under medium human development index with score of above 0.507 and below 0.634. Nine countries fall under high development with HDI of between 0.634 and 0.749. Algeria, Mauritius and Seychelles fall under very high human development with HDI of 0.750 and above. Meanwhile, Algeria had the highest number of death and also one of the highest case fatality rate in Africa.

For SDG score, only 34 countries score above 50 (compared to Denmark with the best score of 85.2). Algeria, Tunisia and Morocco have the best SDG scores of Africa with 71, 70 and 69.1 respectively. Others, including South Africa, Mauritius, Ghana, Gabon, Cape Verde and Sao Tome and Principe have SDG score above 60. Again Egypt, Algeria and Morocco are relatively high on number of death.

Figure 14 shows that some countries (including Algeria, Egypt and Zimbabwe) with good SDG score also have high CFR. Somalia, South Sudan, Central Africa Republic (CAR), Democratic Republic of Congo (DRC), Chad and Sudan are the highest on the GRI with above 7.0. These are followed by Nigeria, Ethiopia, Niger, Mali, Uganda, Mauritania, Kenya, Libya, Mozambique and Burundi with GRI scores above 6.0. Ordinarily, the high GRI countries should have been high on COVID-19 death. However, Fig 14 does not suggest this hypothesis.

**Fig 14:**
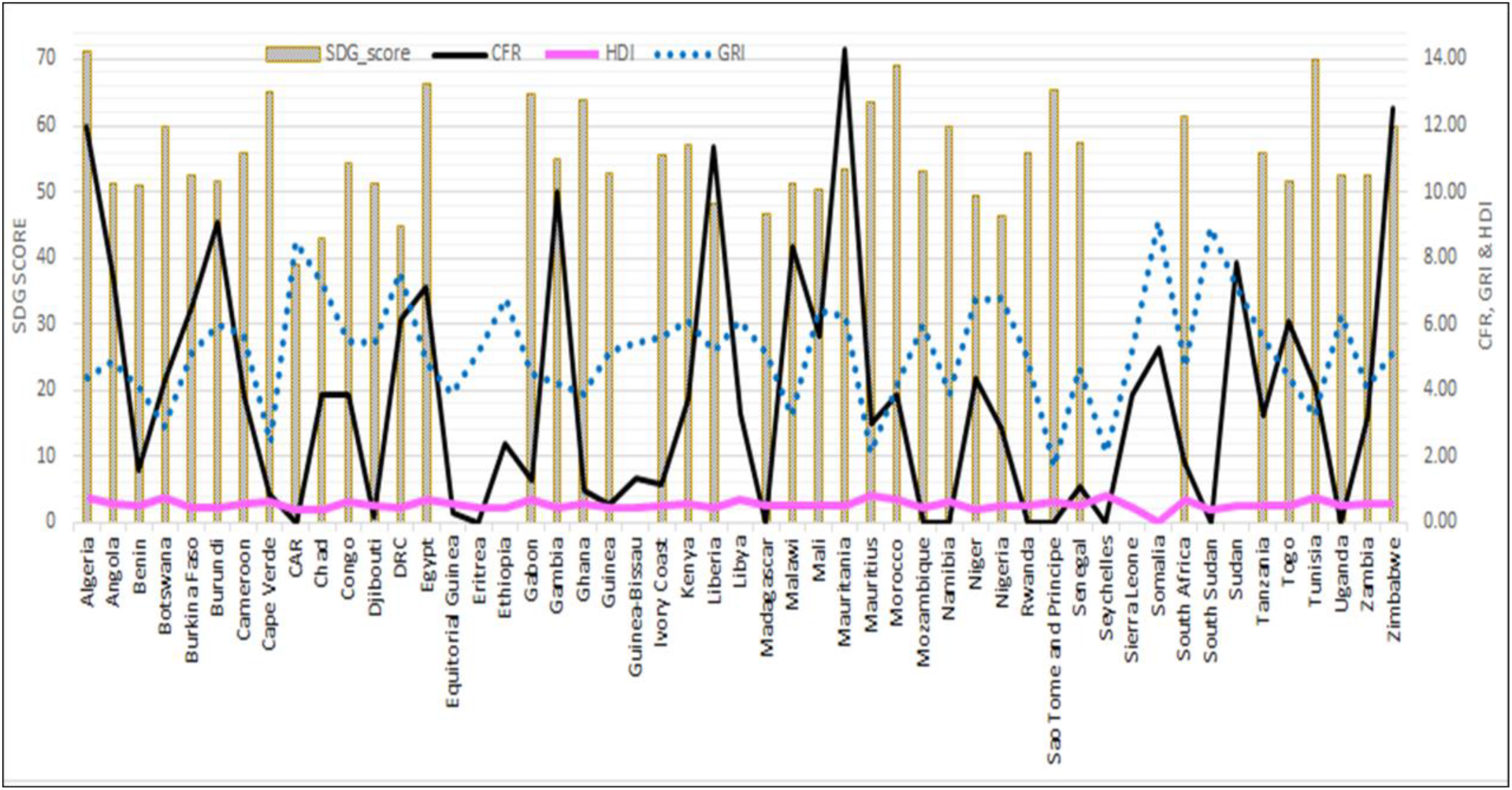
SDG score, HDI, GRI and COVID-19 case fatality rate across Africa.

Although countries may not be able to prevent the incidence of COVID-19, it is expected the more resilience countries (with better performance on HDI, SDG and low GRI) should record less case fatality compared to the less resilient countries. Table 7 shows the Spearman’s correlations between the different variables. Expectedly, Table 7 shows very strong positive correlation among the COVID-19 variables: total test, confirmed cases, and death. However, the correlation with HDI and SDG are weak, with none with the GRI.

**Table 7:** Correlation matrix between SDG score, HDI, GRI and COVID-19.

|  | COVID-19 confirmed | COVID-19 Death | COVID-19 recovery | COVID_ Test | Pop | CFR | Death per 100K | HDI | SDG | GRI |
| --- | --- | --- | --- | --- | --- | --- | --- | --- | --- | --- |
| COVID-19 Confirmed cases | 1 |  |  |  |  |  |  |  |  |  |
| COVID-19 Death | .799** | 1 |  |  |  |  |  |  |  |  |
| COVID-19 Recovery | .917** | .768** | 1 |  |  |  |  |  |  |  |
| COVID-19 Test | .717** | .347* | .672** | 1 |  |  |  |  |  |  |
| Population | .401** | .331* | .290* | .297* | 1 |  |  |  |  |  |
| CFR | .089 | .330* | .116 | -.069 | .055 | 1 |  |  |  |  |
| Death per 100K | .538** | .712** | .598** | .143 | .006 | .297* | 1 |  |  |  |
| HDI | .324* | .314* | .335* | .298* | .003 | -.056 | .382** | 1 |  |  |
| SDG | .340* | .277* | .348* | .241 | .043 | .181 | .327* | .383** | 1 |  |
| GRI | -.076 | -.062 | -.091 | -.120 | .305* | .084 | -.254 | -.705** | -.396** | 1 |

\*\* . Correlation is significant at the 0.01 level (2-tailed).
\* . Correlation is significant at the 0.05 level (2-tailed).

Table 8 shows that Global Risk Index, SDG score and HDI are not strong predictors of COVID-19 confirmed cases and COVID-19 deaths respectively. These variables account for 23% and 19.6% of variances in confirmed COVID-19 cases and COVID-19 deaths respectively across Africa.

**Table 8:** Model summary (Total confirmed Cases and Total Deaths)

| Model | R | R Square | Adjusted R Square | Std. Error of the Estimate | Change Statistics |  |  |  |  |
| --- | --- | --- | --- | --- | --- | --- | --- | --- | --- |
|  |  |  |  |  | R Square Change | F Change | df1 | df2 | Sig. F Change |
| Confirmed Cases | .479<sup>a</sup> | .230 | .181 | 1104.903 | .230 | 4.677 | 3 | 47 | .006 |
| Total Deaths | .443<sup>a</sup> | .196 | .145 | 74.539 | .196 | 3.830 | 3 | 47 | .016 |

a. Predictors: (Constant), GRI, SDG, HDI

## Discussion

In this evaluation, the disease outcomes of the COVID-19 infections among the African countries revealed wide variation in the total number of confirmed cases of the infection. The wide variation in the number of confirmed cases in Africa are attributable to several confounding factors such as international connectivity, climate, healthcare infrastructure, population density, initial responses to the disease outbreaks and national testing capacities. While many of these factors will definitely have some influences on the incidence and prevalence of the disease, the consensus within the scientific community based on views and opinion being expressed in many online platforms is that testing capacity for COVID-19 among the African countries is generally very low, and this would have a major influence on the number of reported confirmed cases of the infection (Otitoloju, 2020). There is little doubt that the total number of confirmed cases of the infection is likely to increase significantly when testing capacities across Africa improves and when rapid antibody-based tests are introduced. The regional comparisons of the total number of confirmed cases also revealed that North Africa is most affected followed by West Africa, Southern Africa, East Africa and Central Africa is least affected. A possible explanation for the distribution is probably the fact that the first cases of the disease was reported in the Northern region especially in Egypt and Algeria before the index cases were reported several days after in other regions (Makoni, 2020). Other possible explanations will be population density and varying testing capacities achievable by the countries in the different regions.

The comparative analysis of confirmed cases of the infection across continents revealed clearly the current level of the disease outcomes is widely varied across continents. Africa and Oceania continents reported very low level of disease outcomes compared to Europe (most affected) followed by North America, Asia and South America. Although the disease outbreak was reported to have started in Wuhan, China, the disease outcomes in Asia were found to be much lower than the levels recorded in Europe and North America. Therefore, the time when index case was reported on the continent does not seem to matter with respect to the eventual magnitude of the prevalence of the disease. The wide variation in reported numbers of COVID-19 infection and deaths across continents also raises a lot of concern about the epidemiology, data collection and management of the COVID-19 on a global scale (Vaughan, 2020). The stark differences in the disease outcomes especially the very high number of recorded deaths due to COVID-19 in countries in Europe and North America compared to countries in Oceania and Africa should necessitate the need for healthcare professionals in the concerned countries to investigate the underlying factors responsible for the wide disparity in COVID-19 disease outcomes across the continents.

In view of the widely reported nature of the COVID-19 infection whereby the majority of people that are infected with the SARS-CoV-2 will be asymptomatic and the wide disparity in testing capacity across Africa, it then suffices that reported death numbers would probably be a more reliable indicator of the disease prevalence among African countries. The results depict that the number of recorded deaths in relation to total confirmed cases was found to be highest in Northern Africa with Algeria having the highest number followed by Egypt and Morocco. Other countries in Africa have recorded relatively low number of deaths during the observation period. On the whole, Africa seems to have recorded a significantly low number of deaths associated with COVID-19 contrary to prediction models which had anticipated up to 300,000 deaths from the coronavirus this year even under the best-case scenario. Under the worst-case scenario with no interventions against the virus, Africa could see 3.3 million deaths and 1.2 billion infections (Gilbert et al., 2020 and UNECA, 2020). The dire consequences of COVID-19 in Africa has been attributed to poor state of health infrastructure, poverty concerns, unfavourable living conditions in the cities, population densities and prevalence of underlying disease conditions like lower respiratory infection, malaria, diarrheal, HIV/AIDS and tuberculosis (The World Bank, 2020). The underlying reasons for the relative low death rates to COVID-19 across African countries when compared to North America, Europe and Asia should warrant further examination of possible factors responsible for this rather unexpected outcome.

Another important outcome of the disease is the recovery cases of COVID-19 in relation to total confirmed cases. Some countries in Africa especially Mauritius, Mauritania, Gambia, Burkina Faso, Uganda, Madagascar and Togo were found to have high recovery rates to the disease. While most of the listed countries are regarded as least developed countries in Africa, rate of recoveries to COVID-19 in these countries has shown that the approaches adopted by some of them may be worthy of emulation by other African countries in response to the COVID-19 pandemic. A cursory look at the management approaches by some of these countries is the strict application of WHO protocol to contain the disease, testing of mainly the people showing the disease symptoms and ensure that all patients who test positive are systematically hospitalised, whether they have symptoms or not, and their contacts are quarantined (www.theafricareport.com). Additionally, most of the countries are providing prompt treatment of the patients with a range of drugs especially Hydroxychloroquine, Chloroquine and Chloroquine-Azithromycin combination (www.africanews.com), despite the current guidance from the World Health Organisation (WHO) that there is no known cure for this virus with 100% certainty. Others have also encouraged the use of natural products with antiviral properties such as COVID-organics in Madagascar and Apivirine (Api-COVID-19) in Burkina Faso.

The outcome of COVID-19 testing capacity across Africa revealed that testing capacity on the continent is very low. Most of the countries on the continent simply have very low capacity to scale up rapidly the capacity to carry out the COVID-19 tests. This low testing capacity relative to population size for COVID-19 across Africa in particular and also around has been widely reported in literature (Oannidis, 2020). At the onset of the outbreak, only two countries in Africa were reported to have the diagnostic capacity for COVID-19. The African Union Commission and the Africa Centres for Disease Control and Prevention (Africa CDC) had launched a new initiative, the Partnership to Accelerate COVID-19 Testing (PACT): Trace, Test & Track (CDC-T3). The partnership is to facilitate implementation of the Africa Joint Continental Strategy for COVID-19, this initiative is said to have been endorsed by African Ministers of Health on 22 February 2020 in Addis Ababa, Ethiopia, and approved by the Bureau of the Assembly of the African Union Heads of State and Government on 26th of March 2020. This initiative is reportedly aimed to strengthen capacity to test for large numbers of COVID-19 across Africa within the next six months (https://africacdc.org/news/). Similarly, the African Union Commission, Africa Centre for Disease Control and Prevention (Africa CDC), and WHO, in partnership with African countries, have established the Africa Taskforce for Coronavirus Preparedness and Response (AFTCOR) working closely to expeditiously scale up diagnostic testing. According to current reports, as of Feb 25, 2020, more than 40 African countries have been empowered to accurately diagnose COVID-19 infection (Nkengasong and Mankoula, 2020).

The factors responsible for the limited number of testing in Africa range from test method adopted or available, laboratory infrastructure, availability of consumables, testing logistics, costs and availability of trained manpower. There are two basic methods adopted for COVID-19 testing – a genetic (RT-PCR) test to detect the virus in a swab sample taken from the nasal and /or throat, and a serological test which detects IgG and IgM antibodies to the virus in blood samples. Currently, the RT PCR molecular diagnostic technique remains the primary method of diagnosing SARs-CoV2 all over the world and indeed on the African continent. Although, the RT-PCR method is a very sensitive method of diagnosis, the method is relatively slow and expensive. This has been a major setback to extensive testing in Africa. Most African countries that could afford the cost of testing are not able to have access to testing kits and consumables due to high demand worldwide. In Africa, strict criteria have been adopted for people under investigations (PUIs) for PCR screening with stringent case definition applied. Other challenges are insufficient number of thermocyclers available and accredited level 3 and 4 Laboratories for testing, and also the low level of manpower required to perform such tests. Most African countries are still in the process of training additional manpower and are also in the process of setting up standard laboratories (Nkengasong and Mankoula, 2020). New test methods are therefore desirable which must be faster, cheaper, safer and produced at a large scale.

It is important to note that some countries in Africa have embraced innovation to solve the problem of testing for COVID-19 in their countries. There are reports of Senegal taking the lead globally in the quest to invent a rapid testing kit for Covid-19. The country is said to have recently developed a rapid test kits that gives COVID-19 test results of an individual in less than 10 minutes at a cost price of one US Dollar (Sh109). This kit is currently undergoing the necessary validation that will allow this test kits for home use. Mass production of the testing kits is proposed to commence immediately in the United Kingdom and Senegal once it meets regulation standards. Reacting to this challenge, Kenya Medical Research Institute has also started manufacturing of Covid-19 testing kits that will give results in fifteen minutes. The kit is reported to be similar to that used in testing HIV with three unilateral lines that will check for any SARS-like virus positivity (https://www.standardmedia.co.ke/).

In this evaluation, no strong relationship currently exists between the global resilient indicators and COVID-19 cases across Africa. Several plausible explanations may be offered for this observation. First, COVID-19 is a new vulnerability that possibly has little regard for existing resilience structures and mechanism. This is supported by the high infection and death incidences recorded even in the so called developed countries in Africa such as Egypt, Algeria, South Africa and Morocco with highly rated resilient systems. Secondly, the observation may also be associated with the variety of initiatives and interventions of multilateral and bilateral agencies that provided several amenities, debt reliefs, grants and loans to many of African countries especially those with lowly rated resilient systems. There is very little doubt that the timely and critical support to developing and low-income countries in Africa from the World Bank, IMF, ADB, Afrexim bank etc has managed to avert what could have been an immediate devastating effects of COVID-19 on the African continent (IMF, 2020). The widespread interventions and initiatives to mitigate the socioeconomic impacts through palliatives measures to reduce the burden of some control methods like lockdown and curfews by African banks, philanthropic organisations and private business leaders. Some private sector organisations such as Coalition Against COVID-19 (CACOVID) in Nigeria helped especially in the provision of extraordinary liquidity assistance and medical facilities and this has contributed significantly to reducing the immediate impact of the disease on the continent. The UN’s efforts which include the health response, coordinated by the World Health Organisation (WHO); the humanitarian response, coordinated by the Office for the Coordination of Humanitarian Affairs (OCHA) and the socio-economic response, coordinated by the UN Development Programme (UNDP) in close collaboration with all UN agencies also contributed immensely to minimizing the impact of COVID-19 pandemic on global resilient indicators in Africa. It is quite possible that the continent might still be in its early days in the fight against the COVID-19 pandemic, so it may not yet be time for complacency. Deliberate efforts must be put in place to ensure vigilance, cooperation, collaboration and identify what measures worked or did not work across the continent. The establishment of channels to share best practices among African countries and develop solutions which take into consideration the peculiarities in Africa must be put in place as we enter into maybe a more exponential phase of the infection or a Post-COVID world.

## Limitation of the study

As the COVID-19 pandemic is still unfolding, this paper has examined the disease outcomes among African countries and carried out cluster analysis based on available data over a 75-day period of observation. The patterns and trends are still evolving and by the time the paper is published, significant changes could have occurred. The wide variation in the number of COVID-19 testing being carried out by the different countries will also have a major impact on the actual number of reported confirmed cases of infection. As the infection is still unfolding, the overall impact of COVID-19 on African resilient indicators will require many years of assessment to finalise its true impacts.

## Conclusion

This study has revealed compelling spatial differences in the incidence, deaths and recoveries from COVID-19 among African countries. The wide variation in the disease outcomes in Africa are attributable to several confounding factors such as international connectivity, climate, healthcare infrastructure, population density, initial responses to the disease outbreaks and differential national testing capacities. The testing capacities for COVID-19 are found to be abysmally low in relation to the population among all African countries. During the 75 days of observation, African countries have recorded significantly low number of deaths associated with COVID-19 contrary to prediction models and comparative death tolls recorded in countries in Europe, North America, South America and Asia. Countries in Africa with highest rate of recovery from the disease were found to have adopted strict adherence to some of WHO protocol to contain the disease, test mainly the people showing the disease symptoms, isolate all those who test positive to the disease and provide prompt treatment of the patients with a range of drugs especially Hydroxychloroquine, Chloroquine and Chloroquine-Azithromycin combination. A few countries of these countries also encouraged the use of natural products with immune-boosting and antiviral properties such as COVID-organics in Madagascar and Apivirine (Api-COVID-19) in Burkina Faso. The study found that no strong relationship currently exists between the global resilient indicators (HDI, SDG and GRI) and COVID-19 cases across Africa. The study recommends that the approaches adopted by some of the African countries which achieved high recovery rates from COVID-19 should be integrated into healthcare management plans for the disease across the continent even as the situation unfolds.

## Data Availability

Yes, data are available on relevant databases.

http://coronavirus.jhu.edu/

http://www.worldometers.info/

## Conflict of Interest

The authors declare that they have no known competing financial interests or personal relationships that could have appeared to influence the work reported in this paper.

## Ethical approval

This study did not involve human or animal experimentation. All data sources are cited and acknowledged throughout the manuscript.

## Acknowledgement

The authors acknowledge the opportunity to serve as volunteers of the UNILAG Consult COVID-19 Advisory Group (UCCAG).

## Funding statement

This research did not receive any specific grant from funding agencies in the public, commercial, or not-for-profit sectors.

## Websites

http://www.africanreview.com/manufacturing/industry/cacovid-orders-supplies-for-400-000-covid-19-tests-kits-for-nigeria

http://www.cacovid.org/

https://africacdc.org/news/african-union-and-africa-centres-for-disease-control-and-prevention-launch-partnership-to-accelerate-covid-19-testing-trace-test-and-track

https://unsdg.un.org/sites/default/files/2020-03/SG-Report-Socio-Economic-Impact-of-Covid19.pdf.

https://www.standardmedia.co.ke/article/2001369392/senegal-develops-one-dollar-rapid-testing-kit-for-covid-19.

https://www.theafricareport.com/26726/coronavirus-recovery-rate-is-there-a-senegalese-exception/

https://www.imf.org/en/News/Articles/2020/04/17/pr20168-world-bank-group-and-imf-mobilize-partners-in-the-fight-against-covid-19-in-africa

www.fda.gov/medical-devices/

www.ncdc.gov.ng

## Notes

### Competing Interest Statement

The authors have declared no competing interest.

### Clinical Trial

Epidemiological data not requiring Clinical Trial ID.

